# Generating complex explanations from machine learning models using class-contrastive reasoning

**DOI:** 10.1101/2023.10.06.23296591

**Authors:** Yujia Yang, Soumya Banerjee

**Affiliations:** Department of Computer Science and Technology, University of Cambridge, Cambridge, United Kingdom

**Keywords:** Explainable AI, complex explanations, class-contrastive analysis

## Abstract

**Objective:** One of the major limitations of most black-box machine learning models is the lack of explainability. In healthcare, explainability is important. Furthermore, most healthcare professionals do not have technical knowledge of machine learning. Consequently, it is necessary to translate the predictions of the machine learning model into an explainable narrative.

Our research focuses on the healthcare domain. The goal of this study is to generate complex explanations from a black-box machine learning model applied to heaalthcare.

**Results:** Class-contrastive techniques can be used to generate explanations. In this method, class-contrastive counterfactual reasoning is applied to a machine learning model on tabular data (in health-care). The model predictions are explained by observing the changes in prediction by altering the inputs. This is visualized using heatmaps (class-contrastive heatmaps). This approach displays prediction results as visualizations (heatmaps).

Our contribution is to extend class-contrastive analysis of black-box machine learning models to numeric features. Our work also allows machine learning scientists to visually inspect class-contrastive heatmaps and generate complex explanations for models. The resulting explanations (visual and text) are easier for non-technical people to follow.

We show how machine learning scientists can extract complex explanations from machine learning models which can be interpreted by nontechnical audiences. Our work may be broadly applicable in domains where explainability is important.

## Introduction

Throughout history, humans have faced challenges in effectively preventing and diagnosing diseases. Artificial Intelligence (AI) and machine learning may help in accurate diagnosis of diseases. However, the explainability of black-box machine learning models remains challenging.

This paper focuses on the explainability of a black-box machine learning model applied to medical data. We build on recent work in class-contrastive analyis [1].

Machine learning models in healthcare face many questions of explainability. In the absence of explainability, the prediction generated by the machine learning model may not be useful.

This work uses class-contrastive analysis to analyze a healthcare dataset. We extend a previous approach [1] to account for numeric features. Our approach explains the model predictions using class-contastive heatmaps. This shows intuitively how each feature affects the outcome.

In addition, we show how machine learning scientists can use these class-contrastive heatmaps to generate complex explanations. These can assist non-technical practitioners (like clinicians and patients) in understanding machine learning models and their results.

## Main text

### Class-contrastive analysis

Compared to a targeted outcome of interest, a class-contrastive explanation explains what caused a model to produce the current output rather than the desired output. An example class-contrastive explanation is: “The patient is predicted to have diabetes because she is elderly. If age was lower, then the prediction would be different.”

In our work, we use class-contrastive analysis to determine which characteristics lead to the diagnosis of diabetes. Heatmaps were created to illustrate the impact of each factor on the risk of diabetes. Using these heatmaps, we were able to generate complex explanations. A previous study has applied class-contrastive reasoning to categorical features and explained machine learning models to predict mortality in patients [1]. Our approach extends work in that study to numeric features.

### Data

This work uses the PIMA dataset from the National Institute of Diabetes and Digestive and Kidney Diseases [2]. It has data on the medical diagnosis of diabetes in women. The dataset is composed of eight medical predictors and one outcome variable (diabetes diagnosis). The features are number of previous pregnancies, blood glucose level, blood pressure (BP), skin thickness (ST), insulin, body mass index (BMI), diabetes pedigree function (DPF) [diabetes likelihood based on patient characteristics] and age [2].

### Implementation details of artificial neural network

We used artificial neural networks to predict the probability of diabetes. The input features are eight characteristics relevant to diabetes, and the predicted output is the probability of diabetes. We used the sequential network from the *tensorflow* and *keras* packages [3] [4].

Our model has three layers. In the first layer, there are 12 neurons with rectified linear unit (ReLu) as their activation function. The second layer consists of eight neurons activated by ReLu. There is one neuron in the last layer whose activation function is the sigmoid function. *Adam* is used as the optimization algorithm.

The ReLu activation function outputs itself if the input is positive; otherwise, it outputs zero. The function is shown below:

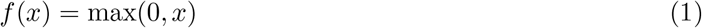

Our model uses the ReLu activation function in the first two layers in an effort to eliminate the problem of vanishing gradients and improve learning.

The problem of predicting diabetes diagnosis is a binary classification task. In the last layer of the model, we used a sigmoid activation function and a binary cross-entropy loss function. The sigmoid function is capable of mapping values to a range between 0 and 1, which corresponds to the range of probability. The function is shown below:

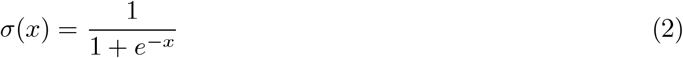

The difference between predicted probabilities and actual class labels is measured using a binary cross-entropy loss function. The function is represented as follows:

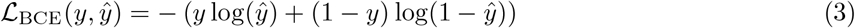

where *y* is the true label (0 or 1) and *ŷ* is the predicted probability of the positive class.

We performed a 50%-25%-25% training-test-validation split of the data. We performed 10-fold cross-validation with regularization to penalize for model complexity.

### Single feature class-contrastive analysis

We first present our class-contrastive analysis on single features.

A single feature in the test set is set to its maximum value and the diabetes diagnosis probability (predicted by the model on the test set) is recorded. Next, the single feature is set to its minimum value and the diabetes probability predicted by the model is recorded. The difference in these probabilities is stored. We repeat this process independently for all features. We then display the difference in probabilities as a heatmap.

In this class-contrastive heatmap with rows representing patients and columns representing features, we visualized the difference in diabetes prediction probability. The heatmap showing the class-contrastive analysis of single features is shown in Fig. 1.

**Figure 1.**
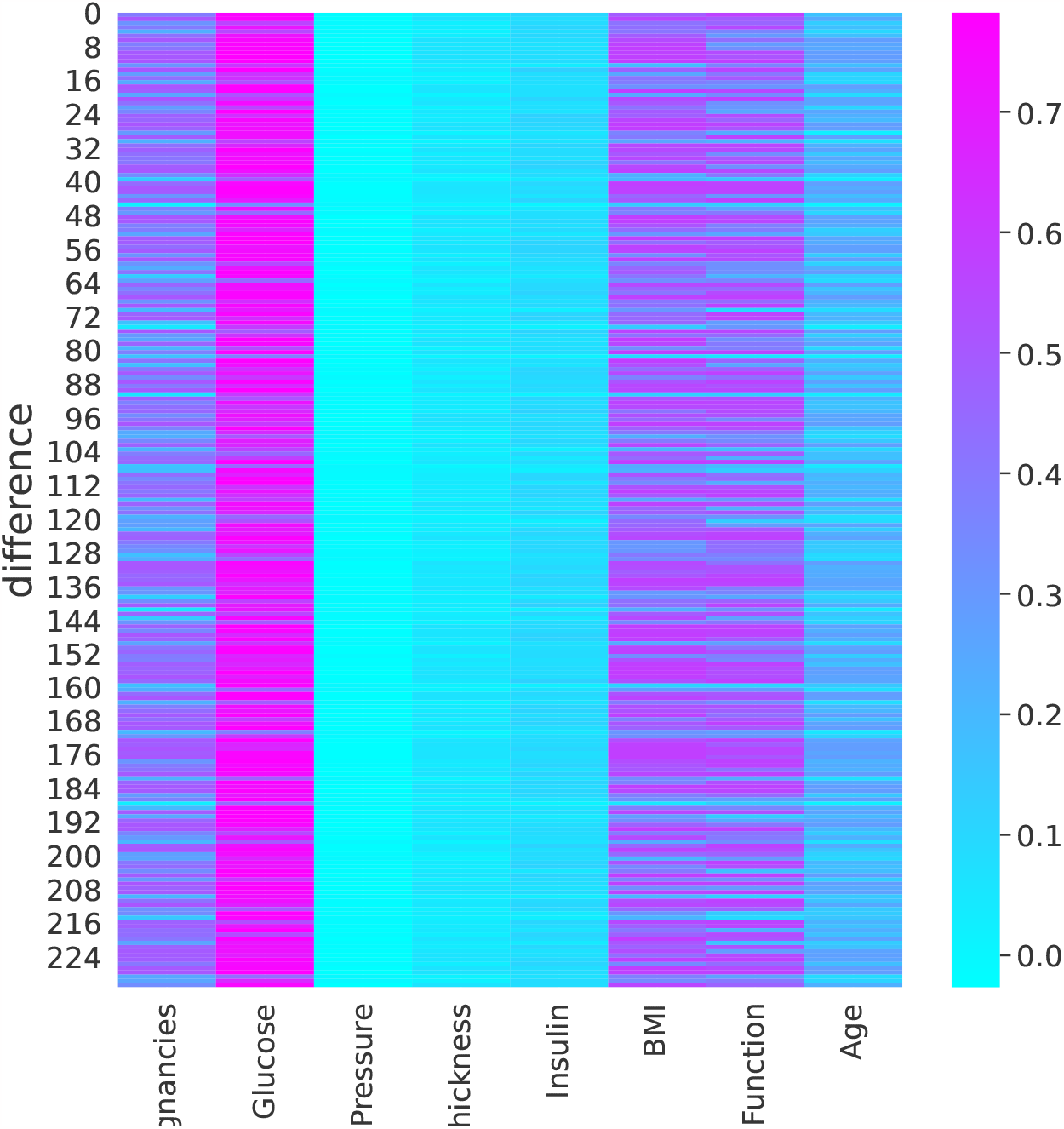
Single feature class contrastive analysis heatmap. Class-contrastive heatmap shows the effect of changing features on machine learning model predictions (on the test set). A single feature is set to its maximum value and diabetes diagnosis probability (predicted by the model on the test set) is recorded. Next, the single feature is set to its minimum value and the diabetes probability predicted by the model (on the test set) is recorded. We repeat this process for all other features. We then display the difference in probabilities as a heatmap. Each row represents a patient and each column represents a feature. Each cell represents the probability difference. The heatmap shows that glucose highly correlates with diabetes diagnosis. High glucose means the patient is at high risk of diabetes. Pregnancy, body mass index [BMI], and diabetes pedigree function [DPF] are also significant factors for diabetes diagnosis. Blood pressure does not seem to predict diabetes diagnosis.

According to the heatmap (Fig. 1), glucose changes are correlated with the diagnosis of diabetes. A high glucose level is indicative of a high risk of diabetes.

Furthermore, pregnancy, body mass index (BMI), and DPF play an important role in the diagnosis of diabetes. The risk of diabetes increases with multiple pregnancies, a high BMI, and a high DPF.

Diagnosis of diabetes is not significantly correlated with skin thickness (ST), insulin, or age. We found that blood pressure is not a reliable indicator of diabetes diagnosis.

### Double feature class-contrastive analysis

In this section, we present results from a double-feature class-contrastive analysis.

Two features in the test set are simultaneously set to their respective maximum values and diabetes diagnosis probability (predicted by the model on the test set) is recorded. Next, the two features are set simultaneously at their minimum values, and the predicted probability of diabetes by the model is recorded. The difference in these probabilities is stored. We repeat this process independently for all combinations of two features. This will yield 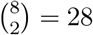 combinations.

We display the difference in probabilities as a heatmap. Patients are represented by rows, and feature combinations are represented by columns (Fig. 2).

**Figure 2.**
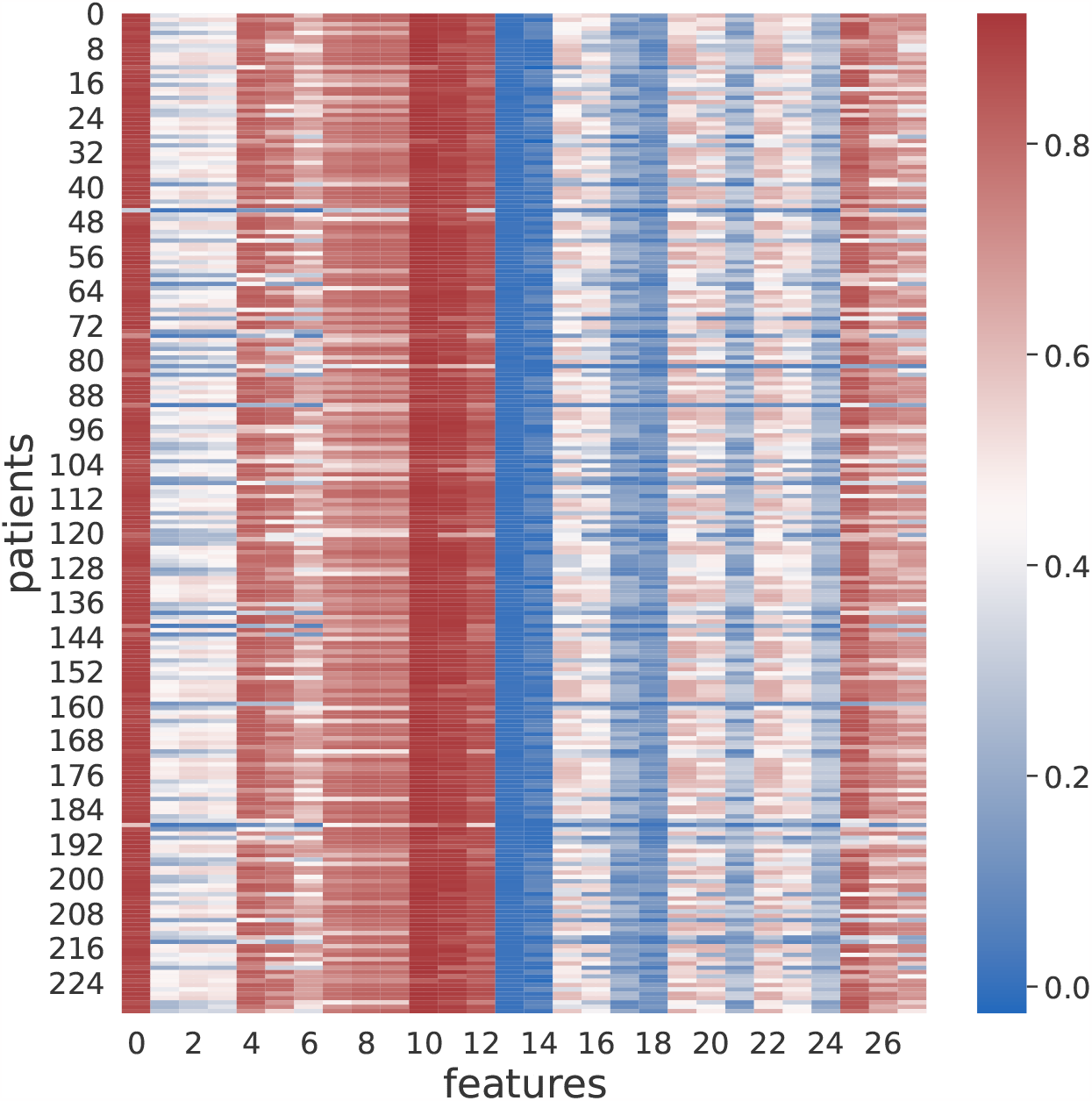
Double feature class-contrastive analysis heatmap. Class-contrastive heatmap showing the effect of changing feature combinations on machine learning models on the test set. Two features were simultaneously set to their respective maximum values and model predicted diabetes probability (on the test set) is recorded. Next, we set the same two features simultaneously to their respective minimum values and recorded the change in diabetes probability. We repeat the process for every combination of two features. We display the difference in probabilities as a heatmap. Each row represents a patient and each column represents a feature. Each cell represents the probability difference. Column 0 (Pregnancy& Glucose), Column 10 (Glucose & body mass index [BMI]), and Column 11 (Glucose & diabetes pedigree function [DPF]) showed a strong correlation with diabetes diagnosis. The higher the value of the two features, the greater the possibility that the patient is diabetic. Column 7 (Glucose & blood pressure [BP]), Column 8 (Glucose & skin thickness [ST]), Column 9 (Glucose & Insulin), and Column 12 (Glucose & Age) were important indicators of diabetes diagnosis. Column 13 (BP & ST) and Column 14 (BP & Insulin) showed a low correlation to diabetes diagnosis.

According to the results of the single feature class-contrastive analysis, glucose changes are the most important factor in the diagnosis of diabetes. Therefore, we would be more interested in the combinations related to glucose in the result of a double feature class-contrastive analysis.

Column 0 (Pregnancy& Glucose), Column 10 (Glucose & body mass index [BMI]), and Column 11 (Glucose & DPF) showed a strong correlation with diabetes diagnosis in Figure 2. The higher the value of the two features, the greater the possibility that the patient is diabetic.

Column 7 (Glucose & blood pressure [BP]), Column 8 (Glucose & ST), Column 9 (Glucose & Insulin), and Column 12 (Glucose & Age) were important indicators for the diagnosis of diabetes. Column 13 (blood pressure [BP] & skin thickness [ST]) and Column 14 (BP & Insulin) showed a low correlation with diabetes diagnosis. This matched the result in the single-feature class-contrastive analysis, where we found that blood pressure did not predict the diagnosis of diabetes.

### Triple feature class-contrastive analysis

A triple feature class-contrastive analysis involves changing three features simultaneously from their maximum to minimum values, respectively.

Three features in the test set are simultaneously set to their respective maximum values and diabetes diagnosis probability (predicted by the model on the test set) is recorded. Next, the three features are set simultaneously to their minimum values, and the probability of diabetes diagnosis (predicted by the model in the test set) is recorded. The difference in these probabilities is stored. We repeat this process independently for all combinations of three features. This will yield 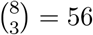 combinations.

We then display the difference in probabilities as a heatmap. Each row represents a patient, and each column represents the three feature combinations we modified (Fig. 3).

**Figure 3.**
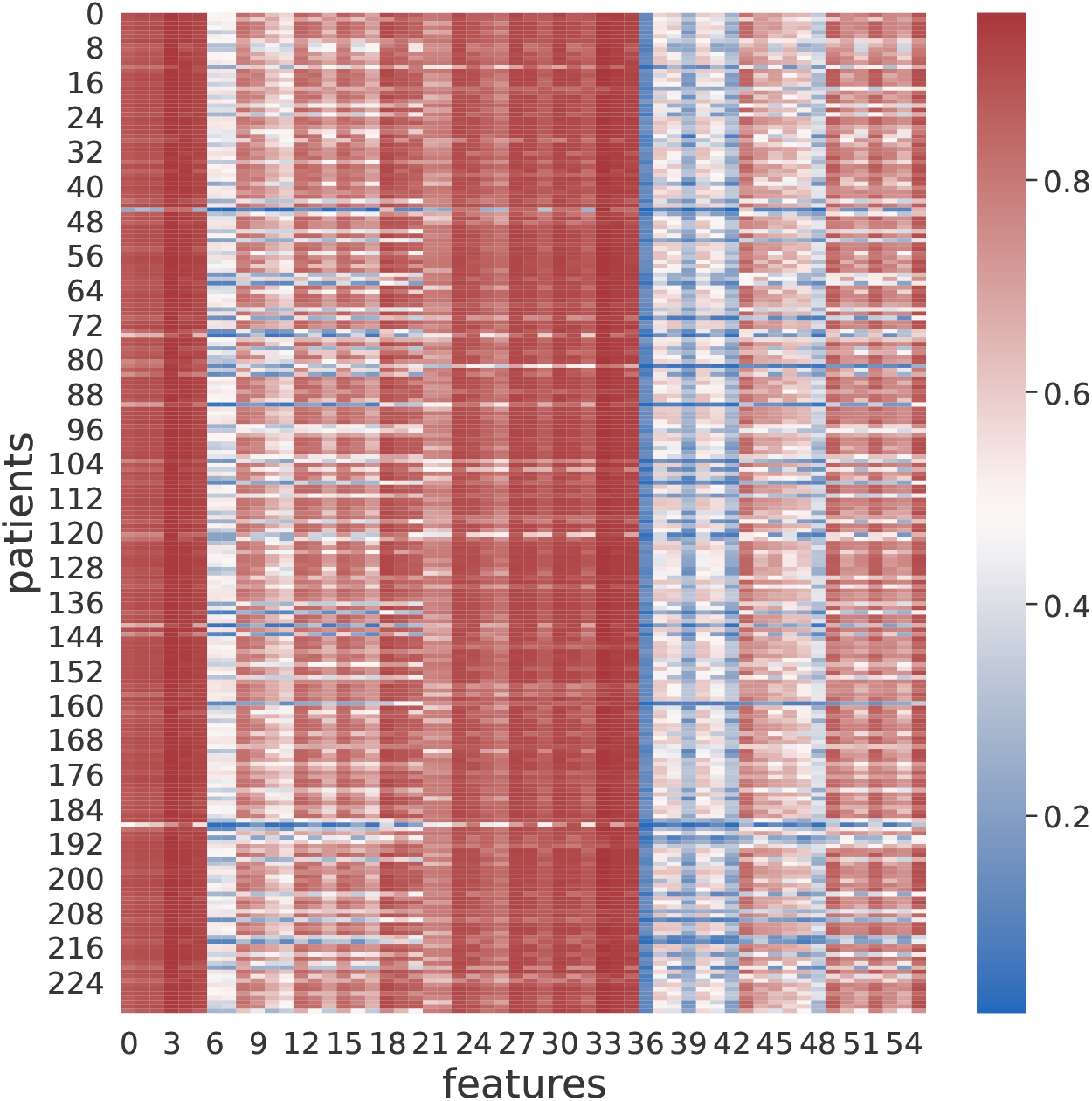
Triple feature class-contrastive analysis heatmap. Class-contrastive heatmap showing the effect of changing 3 features simultaneously on model prediction in the test set. Three features were simultaneously set to their respective maximum values and the probability of diabetes (predicted by the model on the test set is recorded). Next, we set the same three features to their respective minimum values and recorded the diabetes probability predicted by the model. The process is repeated for all combinations of 3 features. We display the difference in probabilities as a heatmap. Each row represents a patient and each column represents a feature combination. Each cell represents the probability difference. The first six columns (index 0 to 5) which represented all the feature combinations with pregnancy and glucose were strongly correlated with diabetes diagnosis. This matched with the result from the double features class-contrastive analysis. Column 6 (Pregnancy & blood pressure & skin thickness), and Column 7 (Pregnancy & blood pressure & insulin) did not affect diabetes diagnosis appreciably.

From left to right, the first six columns (indices 0 to 5) present all combinations of pregnancy and glucose, such as pregnancy & glucose & blood pressure, pregnancy & glucose & skin thickness, pregnancy & glucose & insulin, pregnancy & glucose & BMI, pregnancy & glucose & DPF, and pregnancy & glucose & age. Based on the heatmap, most diabetes probabilities are greater than 0.7, which is indicated by the red colour.

Compared to the other feature combinations, these combinations have a strong correlation with diabetes diagnosis. This also agrees with the results obtained from the double-feature class-contrastive analysis.

Furthermore, the next two columns (columns 6,7) display mainly blue or white colours, indicating that changing pregnancy & blood pressure & skin thickness, and pregnancy & blood pressure & insulin would not affect the diabetes diagnosis probability that much. These results are also consistent with the double feature class-contrastive analysis.

### Clinical implications

Doctors should pay particular attention to changes in glucose levels when limited test results are available. Furthermore, the previous pregnancy history of female patients, the body mass index [BMI value], and the value of the diabetes pedigree function [DPF] should also be monitored together. An increase in both values may result in the onset of diabetes.

In contrast, if the patient has only minor changes in these features or the indicators are within the normal range, there maybe less risk of diabetes developing.

We will later use these results to create complex explanations (see Subsection Complex explanations)

### Complex explanations

Our approach can be used by machine learning scientists to visually inspect the class-contrastive heatmaps and generate complex explanations such as the following.

#### Positive case

> - If the patient has high glucose, multiple pregnancies and is elderly
> - then they maybe at high risk for diabetes.

#### Negative case

> - If the patient has high blood pressure, high skin thickness and high insulin level,
> - then they maybe still at low risk of diabetes.

Such complex explanations, in the form of if-then statements, can be easily explained to nontechnical people (such as patients and clinicians).

## Discussion

The class-contrastive analysis of the combination of features suggests that high glucose levels and multiple pregnancies are important factors for the diagnosis of diabetes. The single-feature class-contrastive analysis suggests that glucose is an important factor for the diagnosis of diabetes. From the double feature class-contrastive analysis we observe that (Pregnancy & Glucose), (Glucose & BMI), and (Glucose & DPF) are important indicators of diabetes diagnosis. The triple feature class-contrastive analysis suggests that feature combinations with pregnancy and glucose (such as pregnancy & glucose & blood pressure, pregnancy & glucose & skin thickness) heavily influence the diabetes diagnosis probability.

The key contribution of our work is to 1) extend the class-contrastive analysis technique to numerical features, and 2) use large combinations of features to generate class-contrastive explanations for machine learning models. This can be used by machine learning scientists to generate complex explanations for machine learning models.

Our complex explanations are similar to stories. Humans use complex stories to communicate with each other [5]. Storytelling and understanding maybe one of the most important differences between humans and other primates [6] [7] [8] [9].

## Limitations

The PIMA dataset contains only eight features. A combinatorial explosion will occur as we add more features and conduct a class-contrastive analysis for each and every combination of these features. In addition, the PIMA dataset only contains tabular data. Datasets with more complex structures, such as images or text, will require a different approach.

Finally, in this work, machine learning experts generated explanations and narratives based on visual inspection of heatmaps. Automatically generating narratives as explanations from machine learning models will be the subject of future work.

## Data Availability

Our software is freely available in the following repository:
https://github.com/neelsoumya/complex_stories_explanations

https://github.com/neelsoumya/complex_stories_explanations

## Abbreviations

DPF: diabetes pedigree function

## Declarations

### Ethics approval and consent to participate

No ethics approval and consent to participate was necessary.

### Consent for publication

Not applicable.

### Availability of data and materials

All software was written in the Python programming language. Our software is freely available in the following repository:

https://github.com/neelsoumya/complex stories explanations

### Author’s contributions

YY carried out the analysis and implementation, participated in the design of the study, and wrote the manuscript. SB designed the study and wrote the manuscript. SB directed the study. All authors gave their final approval for publication.

## Acknowledgements

We acknowledge the help and support of the Accelerate technical team. We are especially grateful to Neil Lawrence and Jessica Montgomery for fruitful discussions and feedback.

## Authors’ information

SB is a Senior Research Fellow at the University of Cambridge. He worked in industry for many years before completing a PhD in applying computational techniques to interdisciplinary topics. He has worked closely with domain experts in finance, healthcare, immunology, virology, and cell biology.

YY is a student at the University of Cambridge and has worked in industry and academia.

